# Investigating antibody reactivity to the intestinal microbiome in severe myalgic encephalomyelitis/chronic fatigue syndrome (ME/CFS)

**DOI:** 10.1101/2023.05.21.23290299

**Authors:** Katharine A. Seton, Marianne Defernez, Andrea Telatin, Sumeet K. Tiwari, George M. Savva, Antonietta Hayhoe, Alistair Noble, Ana Carvalho, Steve James, Amolak Bansal, Thomas Wileman, Simon R. Carding

**Author notes:** Corresponding author: Prof. Simon R. Carding, Food, Microbiome and Health Research Programme, Quadram Institute Bioscience, Norwich Research Park, Norwich, NR4 7UQ, U.K., (e).

## Abstract

Myalgic encephalomyelitis/chronic fatigue syndrome (ME/CFS) is a multisystemic disease of unknown aetiology that is characterised by disabling chronic fatigue and involves both the immune and gastrointestinal (GI) systems. Patients display alterations in GI microbiome with a significant proportion experiencing GI discomfort and pain and elevated blood biomarkers for altered intestinal permeability compared with healthy individuals. To investigate a possible GI origin of ME/CFS we designed a feasibility study to test the hypothesis that ME/CFS pathogenesis is a consequence of increased intestinal permeability that results in microbial translocation and a breakdown in immune tolerance leading to generation of antibodies reactive to indigenous intestinal microbes. Secretory IgA and serum IgG levels and reactivity to intestinal microbes were assessed in five pairs of severe ME/CFS patients and matched same-household healthy controls. For profiling serum IgG we developed IgG-Seq which combines flow-cytometry based bacterial cell sorting and metagenomics to detect mucosal IgG reactivity to the microbiome. We uncovered evidence for immune dysfunction in severe ME/CFS patients that was characterised by reduced capacity and reactivity of serum IgG to stool microbes, irrespective of their source. This study provides the rationale for additional studies in larger cohorts of ME/CFS patients to further explore immune-microbiome interactions.

## Introduction

Myalgic encephalomyelitis/chronic fatigue syndrome (ME/CFS) is characterised by disabling fatigue and autonomic, muscular, cognitive, neurological and immune symptoms that leave patients unable to undertake their pre-morbid work, education, exercise and social activities^1^. A quarter of diagnosed patients are house- or bedbound^2^ and less than 5% ever recover their pre-morbid activity levels^3^. A meta-analysis based on 45 studies estimated an average population prevalence of 0.68% (95%CI = 0.48 to 0.97) ^4^. However, these estimates vary considerably by population and case definition^4^. The prevalence is anticipated to rise following the COVID-19 pandemic as 45.2% of long COVID patients fulfil ME/CFS diagnostic criteria^5^. The most common trigger of ME/CFS is an infection, while other reported triggers include physical or mental trauma and toxin exposure^3^.

Several factors have been implicated in ME/CFS pathogenesis involving the immune (autoimmunity, inflammation and chronic infection), gastrointestinal (GI), neurological, endocrine and metabolic systems^6^. Between 38 and 92% of ME/CFS patients report co-morbid GI disturbances such as IBS^3, 7–9^ and 35% of patients take medication for GI disturbances including pro- and prebiotics, digestive enzymes and sodium bicarbonate^3^. The high co-occurrence of ME/CFS and IBS suggests possible involvement of the intestinal microbiome. Consistent with this possibility several studies have reported changes in the community structure of the stool microbiome of ME/CFS patients exemplified by reduced diversity^8^ and decreased abundance of short-chain fatty acid producing bacterial species^8–12^. ME/CFS patients may also have elevated biomarkers associated with increased intestinal permeability^8, 13, 14^. Intestinal inflammation and increased permeability can compromise immune and microbial tolerance (defined as a state of hypo responsiveness to autologous intestinal microbes) ^15^ leading to hyperreactivity and serum antibody production to autologous intestinal microbes^16^; this can pre-stage autoimmune disease^17^. Of note, ME/CFS patients displaying increased intestinal permeability have a higher incidence of serotonin autoimmunity^18^.

Based upon these observations we hypothesised that ME/CFS pathology is a consequence of the breakdown in immune tolerance resulting from the increased intestinal permeability and microbial translocation that leads to generation of antibodies reactive to autologous intestinal microbes. In support of this proposal, a previous study found that, compared with healthy controls, ME/CFS patients had abnormally high levels of IgA and IgM produced in response to a panel of seven gram-negative enterobacteria^13^. However, this small panel of microbes does not reflect the complexity of the intestinal microbiota which comprises 300-500 bacterial species as well as viruses, archaea and fungi^19^. In addition, IgM antibodies have low specificity for antigens^20^ and IgA is primarily produced at mucosal sites^21^. Serum IgG reactivity to the microbiome in ME/CFS patients has recently been investigated using phage immunoprecipitation sequencing (PhIP-Seq) to screen IgG reactivity to 244,000 bacterial and viral epitopes^22^. This study is, however, restricted to identifying antibody reactivity to peptide antigens and cannot detect reactivity to the immunogenic glycoproteins and lipoproteins that decorate the outer membrane and surface of bacterial cells, viruses and fungi^23^. Furthermore, none of the approaches used to date provide information on whether immune tolerance to indigenous intestinal microbes is lost in ME/CFS.

To begin to address this important question and assess the systemic humoral immune response to indigenous microbes we adapted ‘IgA-Seq’, a method that combines flow-cytometry-based bacterial cell sorting and 16S rRNA sequencing to detect mucosal IgA reactivity to the microbiome^24^, to characterise serum IgG reactivity to the microbiome; we refer to this new method as IgG-Seq. To evaluate the IgG-Seq protocol, we designed a feasibility study with a small cohort of severe ME/CFS patients and healthy controls from the same households. The difficulty in accessing housebound or bedbound patients is a major obstacle to understanding the pathophysiology and aetiology of ME/CFS and is why only 0.5% of ME/CFS research is undertaken in severely affected patients^2^. One aim of this study, therefore, was to assess the feasibility of, and barriers to, including severe ME/CFS patients in research involving the collection of biological samples.

## Results

### Recruiting severe ME/CFS patients and same household controls

Study participants were recruited from the CFS clinic at Epsom and St Helier University Hospitals (ESTH), Carshalton, UK and the ME/CFS service at East Coast Community Healthcare Centre, Lowestoft, UK which together had 3,812 registered patients. Recruitment began in October 2017, with the intention of recruiting ten severe house- or bedbound ME/CFS patients and ceased in April 2020 due to the COVID-19 pandemic. Thirty-six patients were invited to the study with a response rate of 58.3% (Supplementary Fig. S1). Of those who responded, 42.9% were ineligible due to either the absence of a household control (n = 4), failure to meet other inclusion criteria (n = 4) or were unable to provide written informed consent (n = 1). Of the 12 eligible pairs of participants, six provided informed consent. The consenting appointments for the other six eligible participants were delayed either due to patients rescheduling their appointments when feeling unwell, or the non-availability of phlebotomists to attend home visits. Consequently, consenting appointments were delayed by up to 12 months for participants, by which point three patients either saw their health further deteriorate preventing them from participating in the study, or acquired an additional health complication that excluded them from the study. We received no further communication from the remaining three pairs.

### Study population characteristics

Samples were collected from five pairs of participants; one pair consented immediately prior to the COVID-19 pandemic which prevented us from obtaining their samples (Table 1). The recruited patients comprised four females and one male (mean age 33.8 years; standard deviation (SD) 13.8). There was clinical heterogeneity amongst patients, with variation in the age at which ME/CFS onset occurred, length of illness and symptom severity. Three patients reported ME/CFS onset following a viral infection, one following vaccination and one following surgery. IBS was reported in all patients but in none of their matched household controls. Same household healthy controls included four males and one female (mean age 40.4 years; SD 16.7) who were the carers and spouse (n = 3), parent (n = 1) or sibling (n = 1) of the patients.

**Table 1.** Severe ME/CFS clinical characteristics

|  | Participants<br>affected (%) | Mean (SD) | Range |
| --- | --- | --- | --- |
| **Age of ME/CFS onset (years)** | - | 25.0 (9.34) | 12-38 |
| **Length of ME/CFS (years)** | - | 8.4 (6.83) | 2-21 |
| **Symptoms** |  |  |  |
| Post exertional malaise | 100 | - | - |
| Non-restorative sleep | 100 | - | - |
| Headaches of a new onset, pattern and severity | 80 | - | - |
| Recurrent sore throat with enlarged glands in neck | 40 | - | - |
| Impaired concentration | 100 | - | - |
| Impaired memory | 80 | - | - |
| Joint pain | 60 | - | - |
| Muscle pain | 80 | - | - |
| Visual and/or auditory hypersensitivity | 100 | - | - |
| Irritable bowel syndrome | 100 | - | - |
| **Questionnaire (maximum score)** |  |  |  |
| Shortened SF-36 (30) * | - | 11.0 (1.73) | 10-14 |
| Chalder fatigue – physical (28) * | - | 24.0 (3.67) | 18-27 |
| Chalder fatigue – mental (16) * | - | 13.5 (1.12) | 12-15 |
| HADS – anxiety (21) * | - | 7.8 (4.55) | 3-15 |
| HADS – depression (21) * | - | 6.5 (5.50) | 3-16 |
| Self efficacy (60) ** | - | 12.0 (7.35) | 3-21 |
| Visual analogue (100) * | - | 62.5 (36.31) | 0-90 |
| Epworth sleepiness (24) *** | - | 7.5 (14) | 1-14 |

\* questionnaire completed by 4 patients
\*\* questionnaire completed by 3 patients
\*\*\* questionnaire completed by 2 patients

### Stool consistency does not separate severe ME/CFS patients with IBS from matched household controls without IBS

Despite all ME/CFS patients, and no household controls, reporting IBS there was no evidence for a difference in consistency of the collected stool samples between patients and controls, as measured by the Bristol stool form scale (BSFS) or by water content (Fig. 1A and 1B). Three controls had abnormally loose stools (BSFS 5-7) indicative of diarrhoea and one control had an abnormally hard stool (BSFS 2) indicative of constipation. In contrast, two patients had stool samples defined as having a healthy consistency (BSFS 3-4), one patient had an abnormally loose stool (BSFS 5) and two patients had abnormally hard stools (BSFS 1-2).

**FIGURE 1.**
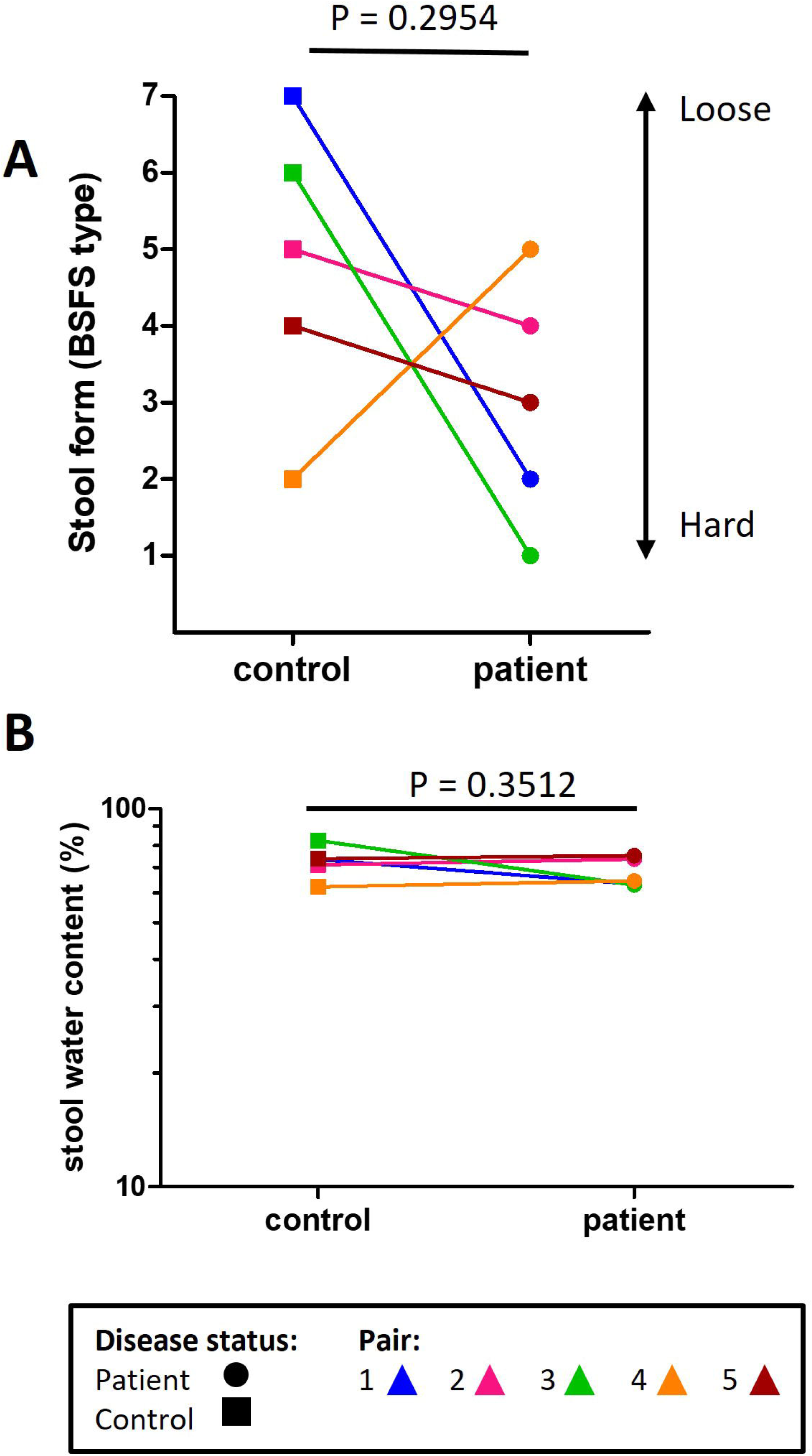
Stool consistency. (A) Analysis of stool consistency using the Bristol stool form scale (BSFS) in severe ME/CFS patients (n=5) and matched household controls (n=5). P values were calculated using a two-tailed paired t-test. (B) Water content in stool samples in severe ME/CFS patients (n=5) and matched household controls (n=5). P values were calculated using a two-tailed paired t-test.

### Assessment of secretory IgA in stool

The concentrations of microbe bound IgA1/2 (Fig. 2A) and free IgA1/2 (Fig. 2B) were measured by ELISA with no evidence of a significant difference between severe ME/CFS patients and their matched household controls. Flow cytometry was used to determine the distribution of IgA coating on stool microbes (Fig. 3A and 3B). When comparing the proportion of stool microbes coated by IgA there was no evidence of any differences between patients and matched controls (Fig. 3C). Based on the microbial load of stool samples (Supplementary Fig. S2) the relative quantification of IgA bound microbes was converted to absolute values which again revealed no evidence of differences between patients and matched controls in the quantity of IgA coated microbes within stool samples (Fig. 3D).

**FIGURE 2.**
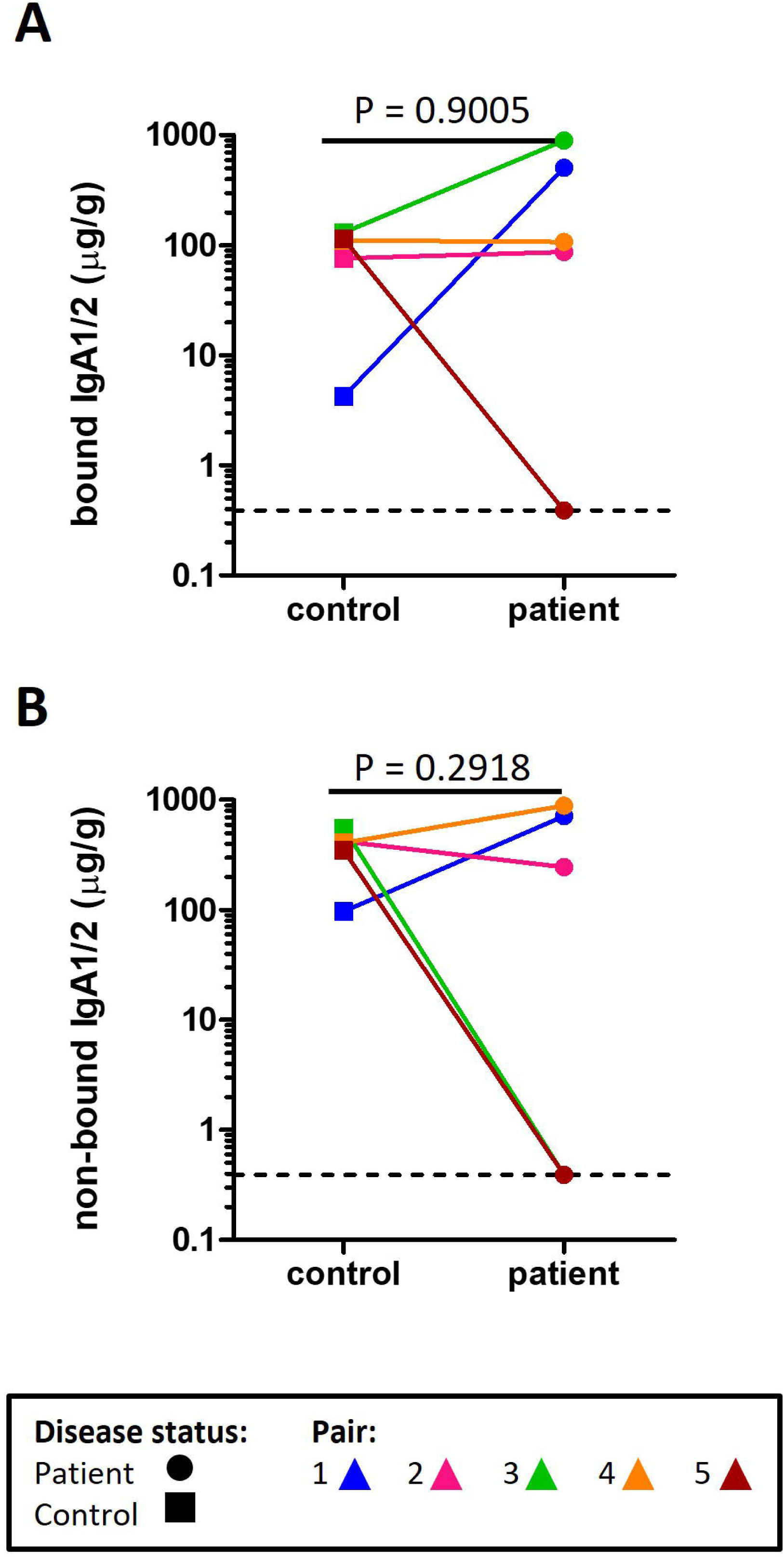
Concentration of IgA measured by ELISA in the stool of severe ME/CFS patients (n=5) and matched household controls (n=5) for (A) microbe bound IgA, and (B) microbe non-bound IgA. For both plots *p values* were calculated using a two-tailed paired *t*-test.

**FIGURE 3.**
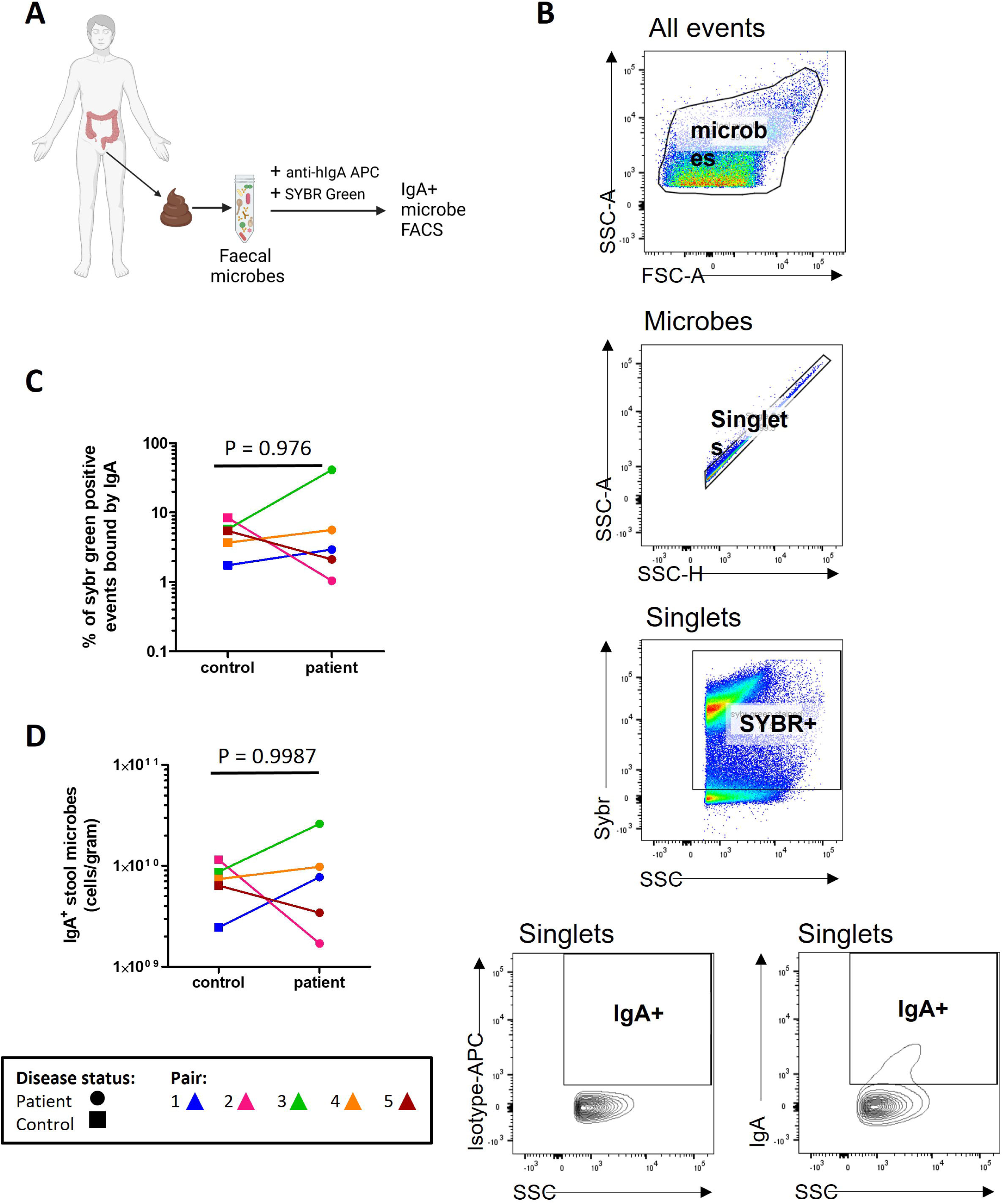
Profiling stool IgA. (A) Overview of sample preparation for IgA-bound microbe fluorescent activated cell sorting (FACS) analysis. (B) Representative flow cytometric analysis of IgA-bound stool microbes. (C) Proportion of stool microbes bound by IgA in severe ME/CFS patients (n=5) and matched household controls (n=5). P values were calculated using a two-tailed paired t-test. (D) Analysis of IgA-bound microbial load in severe ME/CFS patients (n=5) and matched household controls (n=5). P values were calculated using a two-tailed paired t-test.

### Severe ME/CFS patients have a reduced serum IgG immune response to stool microbes

To determine whether immune tolerance to intestinal microbes was altered in severe ME/CFS patients the level of serum IgG antibodies bound to both their own (autologous) stool microbes and to non-self (heterologous) stool microbes from other individuals were measured in patients and their matched household controls (Fig. 4A). Four patients had lower serum IgG reactivity to indigenous stool microbes than their matched household controls (p = 0.07322) and all patients had lower serum IgG reactivity to foreign stool microbes than their matched household controls (p = 0.006334) (Fig. 4B). Furthermore, there was evidence for controls having higher serum IgG reactivity to heterologous (patient) stool microbes than serum IgG reactivity to autologous stool microbes (p = 0.0317). In contrast, for patients there was no evidence for a difference in serum IgG reactivity to autologous and heterologous (control) stool microbes (p = 0.3619). In addition, serum IgG reactivity to patients (heterologous) stool microbes in control individuals was higher than serum IgG reactivity to autologous stool microbes in patients (p = 0.003751). A paired t-test of the averaged IgG levels to indigenous and foreign stool microbes within individuals suggests that patients had a lower IgG reactivity to stool microbes than controls irrespective of the source of the microbes (p = 0.0185). Of note, the reduced levels of patient IgG antibodies to indigenous and foreign stool microbes were not due to lower levels of serum IgG in patients (Fig. 4C).

**FIGURE 4.**
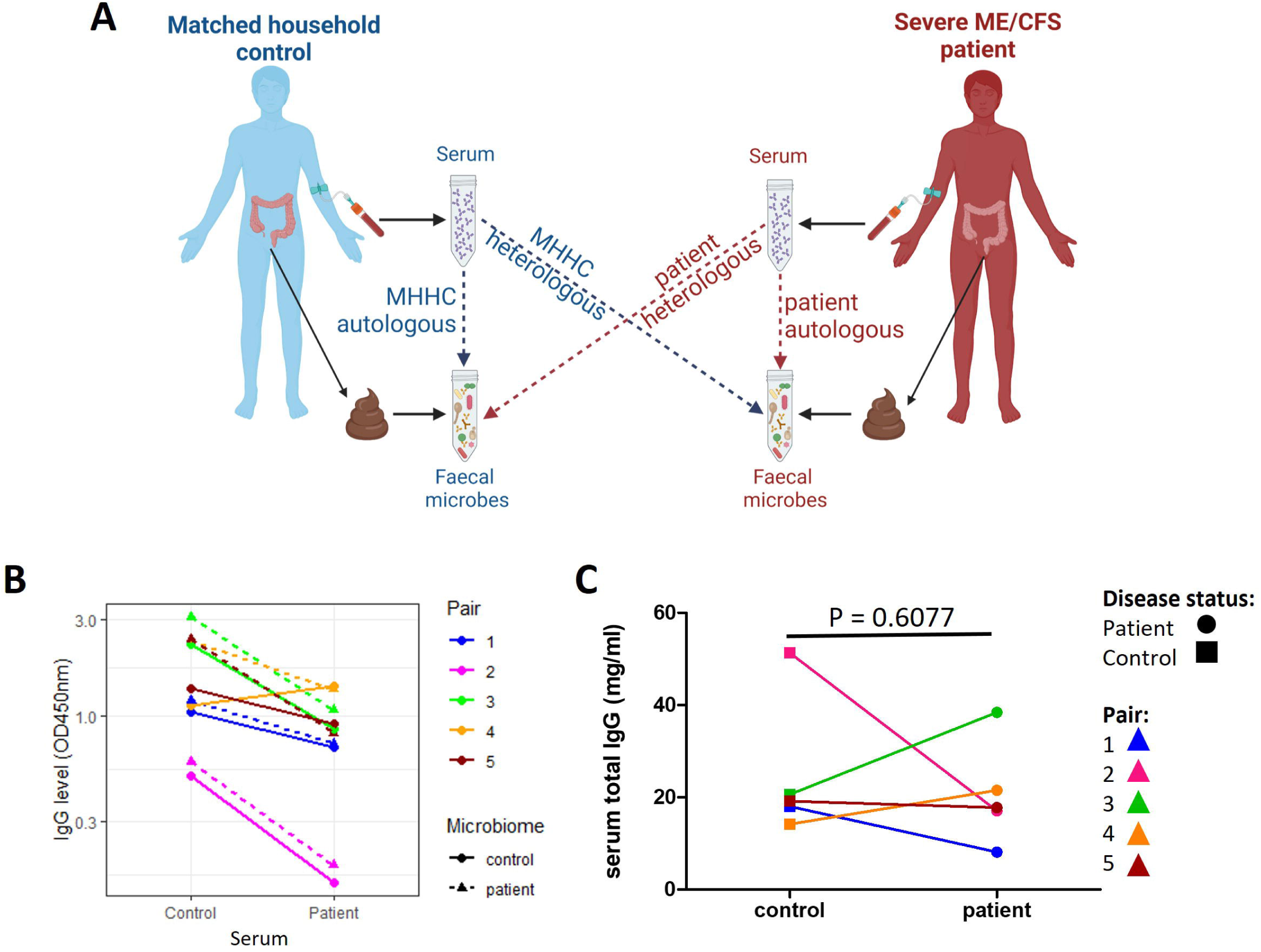
Serum IgG reactivity to autologous and heterologous stool microbes. (A) Overview of assessment of IgG responses to indigenous (autologous) and foreign (heterologous) stool microbes in severe ME/CFS patients and matched healthy household controls. (B) Level of IgG in serum of severe ME/CFS patients (n=5) and household control (n=5) binding to stool microbes from patient and control stool samples in vitro. (C) Levels of IgG in serum of severe ME/CFS patients (n=5) and their matched household controls quantified by ELISA (n=5). Two-tailed paired t-tests were used to calculate P values.

### The proportion of microbes bound by serum IgG is high in both patients and controls

We next sought to determine how much of the stool microbiome was recognised by serum IgG by measuring the proportion of indigenous stool microbes coated by serum IgG (Fig. 5A and 6B). There was no evidence for any differences in the proportion of stool microbes coated by IgG (‘IgG positive’) in severe ME/CFS patients compared with controls (Fig. 5C).

**FIGURE 5.**
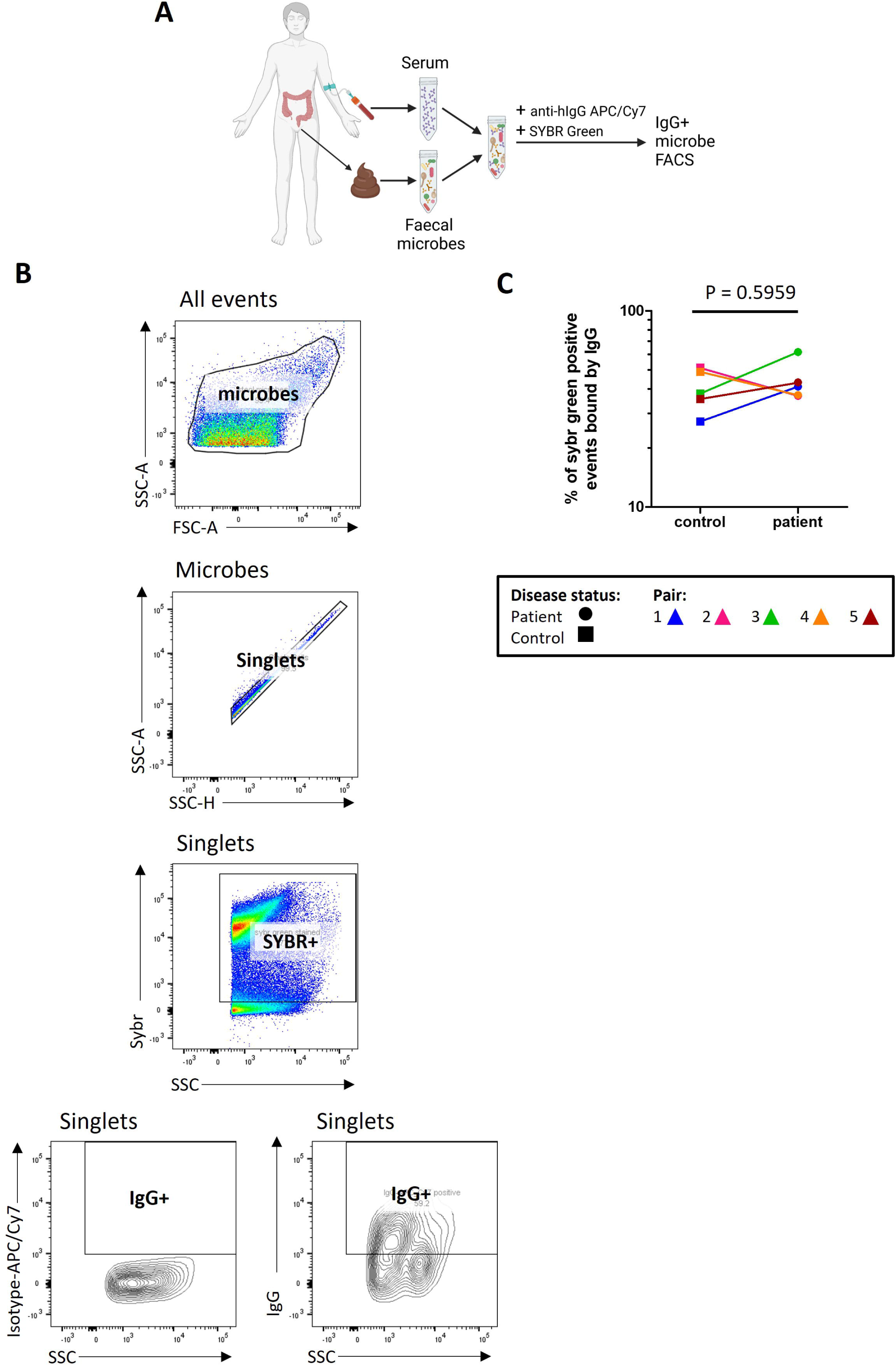
Quantifying stool microbes recognised by autologous serum IgG. (A) Overview of sample preparation for IgG-bound microbe FACS analysis. (B) Representative analysis of serum IgG binding to stool microbes. (C) Analysis of the proportion of stool microbes bound by serum IgG in severe ME/CFS patients (n=5) and matched household controls (n=5). P values were calculated using a two-tailed paired t-test.

**FIGURE 6.**
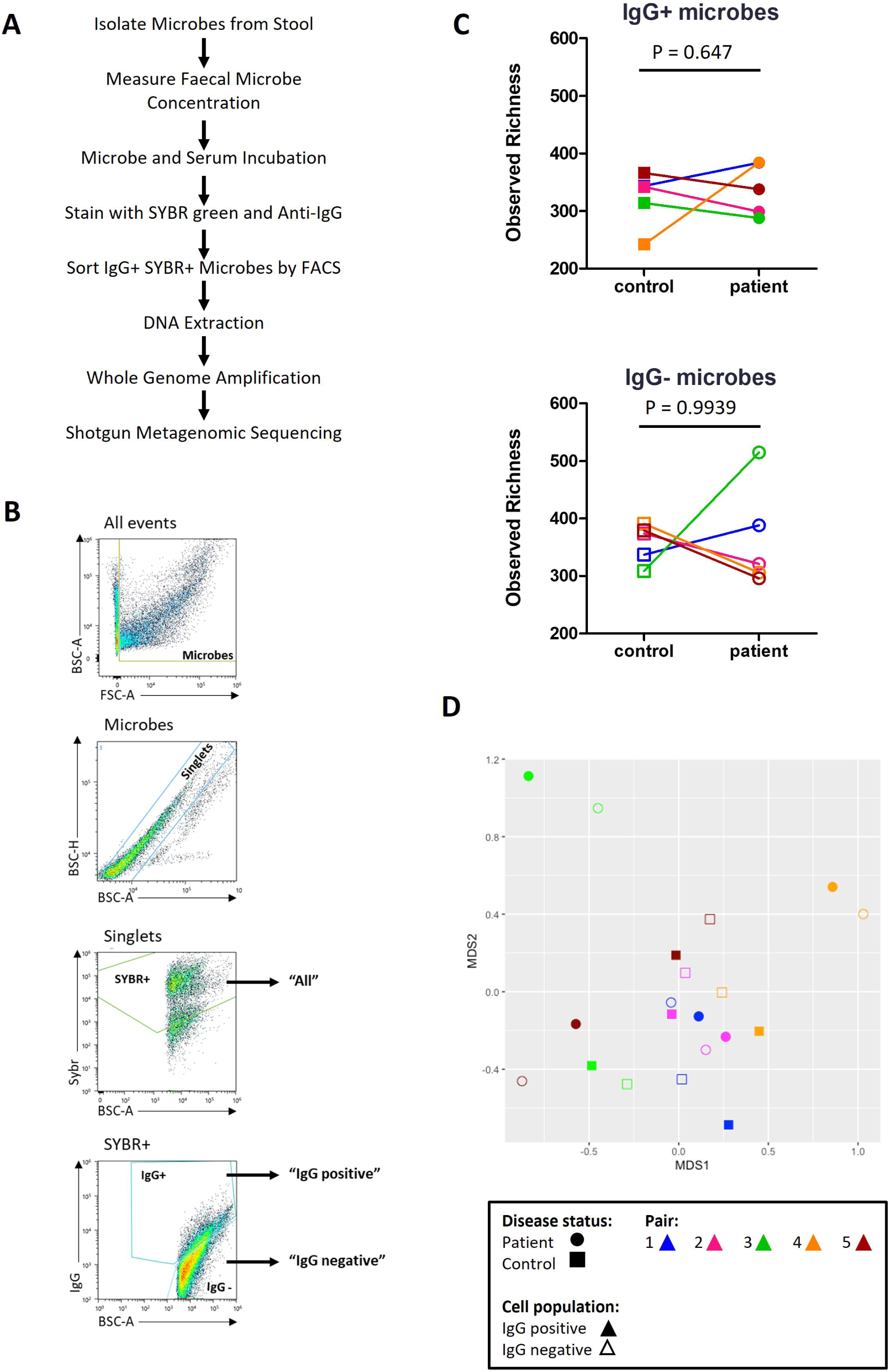
IgG-Seq. (A) IgG-Seq protocol used to determine taxa from stool samples reactive to autologous serum IgG. (B) Representative flow cytometric dot plot showing IgG-Seq gating strategy based upon forward scatter (FSC) and back scatter (BSC) characteristics, followed by discrimination of singlets from doublets. SYBR+ microbes were discriminated from auto fluorescent debris and collected for profiling of all stool microbes. IgG positive and IgG negative populations were sorted from SYBR+ events. (C) Pairwise comparisons of observed richness of IgG positive and IgG negative microbes in severe ME/CFS patients (n=5) and matched household controls (n=5). Analyses were performed at the species-level reads rarefied to the lowest sequencing depth. P values were measured using two-tailed paired t-tests. (D) Beta diversity of IgG positive species (filled shapes) and IgG negative species (unfilled shapes) in severe ME/CFS patients (circles) and household controls (squares). Beta-diversity was calculated using Bray-Curtis dissimilarity, presented on a non-metric multi-dimensional scaling (NMDS) plot.

### Characterising the stool microbiome

Using whole metagenome shotgun sequencing the microbial composition of SYBR Green^+^ (‘all’) stool microbes isolated by fluorescent activated cell sorting (FACS) was determined (Supplementary Fig. S3). The average number of ‘all’ microbes collected by FACS was 1.47 million. Taxa with a relative abundance greater than 1 x 10^-6^ were included in downstream analyses and comparisons made at the genus and species level.

At the genus level 275 taxa were detected. The 15 most abundant genera across all ME/CFS and matched household control samples (n = 10) were *Bacteroides* (10.4%), *Phocaeicola* (10.1%), *Clostridioides* (9.5%), *Lysobacter* (9.4%), *Faecalibacterium* (8.6%), *Blautia* (7.1%), *Roseburia* (5.8%), *Anaerostipes* (5.5%), *Akkermansia* (4.4%), *Campylobacter* (2.9%), *Agrobacterium* (2.6%), *Methanobrevibacter* (1.9%), *Bifidobacterium* (1.9%) *Anaerobutyricum* (1.5%) and *Streptococcus* (1.3%) (Supplementary Fig. S3A).

At the species level 705 taxa were detected. The 15 most abundant species across all ME/CFS and matched household control samples (n = 10) were *Clostridioides difficile* (9.5%), *Lysobacter enzymogenes* (9.4%), *Faecalibacterium prausnitzii* (8.6%), *Phocaeicola dorei* (6.4%), *Blautia* sp. SC05B48 (6.3%), *Anaerostipes hadrus* (5.4%), *Roseburia intestinalis* (4.5%), *Akkermansia muciniphila* (4.3%), *Bacteroides uniformis* (3.7%), *Phocaeicola vulgatus* (3.6%), *Agrobacterium tumefaciens* (2.5%), *Methanobrevibacter smithii* (1.8%), *Bacteroides cellulosilyticus* (1.6%), *Anaerobutyricum hallii* (1.5%) and *Campylobacter jejuni* (1.5%) (Supplementary Fig. S3C).

Using three measures of intra-sample diversity, the Shannon index, inverse Simpson index and observed richness, there was no evidence for differences in any of the alpha diversity measures between severe ME/CFS patients and matched household controls at the species level (Supplementary Fig. 4A).

Due to the large variation in microbial load in stool samples from severe ME/CFS patients (6.3 x 10^10^ cells/gram to 2.6 x 10^11^ cells/gram) and controls (1.2 x 10^11^ cells/gram to 2.0 x 10^11^ cells/gram) (Supplementary Fig. S2), converting relative abundances to absolute abundances increased the heterogeneity amongst samples at both the genus and species levels (Supplementary Fig. S3B and S2D). However, analysis of beta diversity using Bray-Curtis dissimilarity identified subtle changes in both relative microbiome profiles (RMP) and quantitative microbiome profiles (QMP). Samples from patients in pairs three, four and five were most dissimilar to the other samples while samples from patients in pairs one and two clustering together with their matched household controls (Supplementary Fig. S4B).

Functional differences in the microbiome of patients and controls were determined by comparing the abundance of gene families in the ‘all’ fraction. A total of 464,263 gene families were detected in all participants above the threshold. A further filtering step removed gene families below the threshold in more than seven samples, leaving 84,888 gene families for evaluation. PCA was used to reduce the number of variables by defining principal components (PC) that highlighted the largest sources of variation amongst the samples. PC4 identified 11% of variation in functional genes families amongst samples that were attributable to disease status (Supplementary Fig. S5).

### IgG-Seq identifies antimicrobial signatures unique to each participant

Next, IgG-Seq was used to characterise the indigenous gut microbes bound by serum IgG in severe ME/CFS patients and matched household controls. Briefly, ‘IgG positive’ and ‘IgG negative’ microbes were isolated by FACS from bulk stool samples and processed for shotgun metagenomic sequencing to identify taxa preferentially bound by serum IgG (Fig. 6A and 6B). The mean number of ‘IgG positive’ microbes collected by FACS was 1.05 x 10^6^. The mean number of ‘IgG negative’ microbes isolated by FACS was 1.30 x 10^6^. Taxa with a relative abundance greater than 1 x 10^-5^ in the ‘all’ fraction were included in the ‘IgG positive’ and ‘IgG negative’ fractions for downstream analyses. Taxonomic comparisons were made at the species level.

Observed richness scores were applied to rarefied reads to determine the number of microbial species with serum IgG reactivity (Fig. 6C). All participants had more than 200 species in the ‘IgG positive’ fraction with a high number of species also identified in the ‘IgG negative’ fraction. Both patients and controls had a higher mean number of species in their ‘IgG negative’ fraction than their ‘IgG positive’ fraction (365 vs 339 in patients and 358 vs 322 in controls). However, most participants had a small number of species (<30) exclusively in the ‘IgG negative’ fraction at a relative abundance >1 x 10^-5^, except for the patient from pair three who had 107 species exclusively in their ‘IgG negative’ fraction (Table 2). In addition, this patient also had a smaller proportion of species recognised by IgG compared with all other participants (Table 2).

**Table 2.** Proportion of species detected that are recognised by serum IgG

| Pair | Participant | Number of species detected per fraction |  |  | Proportion of species IgG+ |
| --- | --- | --- | --- | --- | --- |
|  |  | Only IgG- | Only IgG+ | Both IgG- and IgG+ |  |
| 1 | Patient | 7 | 7 | 201 | 0.967 |
|  | Control | 23 | 7 | 173 | 0.887 |
| 2 | Patient | 18 | 7 | 190 | 0.916 |
|  | Control | 21 | 6 | 202 | 0.908 |
| 3 | Patient | 107 | 4 | 154 | 0.596 |
|  | Control | 14 | 12 | 145 | 0.918 |
| 4 | Patient | 4 | 19 | 138 | 0.975 |
|  | Control | 29 | 6 | 168 | 0.857 |
| 5 | Patient | 9 | 18 | 162 | 0.952 |
|  | Control | 15 | 31 | 206 | 0.940 |

Next, we assessed the similarity/difference between ‘IgG positive’ and ‘IgG negative’ fractions in patients and controls using the Bray-Curtis index and visualising distances using an NMDS plot (Fig. 6D). ‘IgG positive’ and ‘IgG negative’ fractions from the same participant shared the greatest similarity. No clustering was seen for either fraction in patients or controls. The ‘IgG positive’ and ‘IgG negative’ fractions from patients in pairs three and five were most dissimilar to the other samples.

Using the probability ratio developed by Jackson et al., (2021)^25^ we scored IgG binding by directly measuring the likelihood of a species being bound by IgG. Positive IgG probability ratios indicate that a species is more likely to be coated with IgG and reside in the ‘IgG positive’ fraction. Conversely, negative IgG probability ratios indicate that a species is more likely to be uncoated and reside in the ‘IgG negative’ fraction. Probability ratios are not influenced by the relative abundance of a species within the ‘all’ fraction and therefore measure the quantity of IgG produced against a given species. Of the 423 species detected at a relative abundance greater than 10^-5^, 101 species were detected in every participant and therefore used in downstream analysis. It is worth noting that despite detecting species falling into the bacteria, fungi, archaea and virus kingdoms, only bacterial species were detected in every participant and consequently used in downstream analysis. Notably, owing to the way in which probability ratios are calculated they tend to be similar across species within participants, with some having consistently lower estimates across all species than others, driven by their overall IgG probability ratio. Probability ratio scores varied amongst species and participants. Each participant had a unique combination of probability ratio scores for different species (Fig. 7). We were unable to meaningfully test for differences in probability ratio scores between patients and controls for individual species due to the small sample size and lack of power to detect significant changes.

**FIGURE 7.**
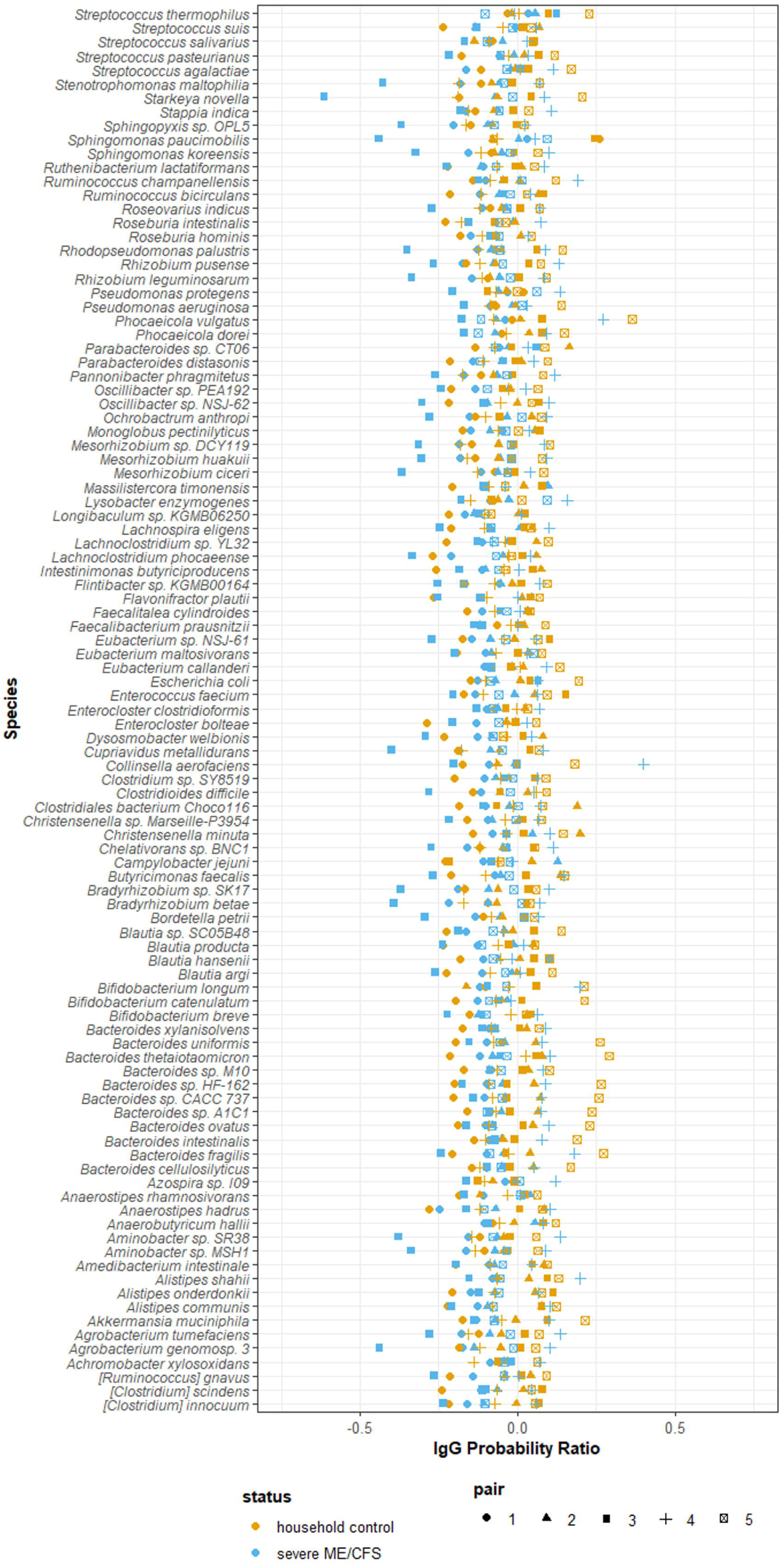
Probability of serum IgG binding to autologous stool microbes from severe ME/CFS patients (n=5) and matched household controls (n=5). IgG probability ratios for species detected in all participants stool samples.

Finally, we analysed the function of the microbial communities within the ‘IgG positive’ and ‘IgG negative’ fractions by analysing the likelihood of a gene family being present in a microbe bound by IgG. IgG probability ratios were calculated for the 1724 gene families that were detected in all participants above the threshold. Using a PCA plot to assess how the ten samples varied with regards to the likelihood of certain gene families being present in microbes bound by IgG (Supplementary Fig. S6), a separation between severe ME/CFS patients and controls from four households was observed (PC2), corresponding to 13% of explained variance.

## Discussion

This feasibility study highlights the logistical challenges of including severe, house- or bedbound ME/CFS patients in research studies and collecting biological samples. Contrary to our initial hypothesis, our findings from a small cohort of patients and controls suggests that severe ME/CFS patients may have a reduced serum IgG immune response to stool microbes.

### Lessons learnt for the inclusion of severe ME/CFS patients in biomedical research

Despite their significant disability severe ME/CFS patients were enthusiastic and eager to participate in our study with a 58% response rate. However, severe ME/CFS often coincided with additional illness complications that excluded these individuals from the study, hindering recruitment. We also found that symptom severity of house- or bedbound ME/CFS patients fluctuated, making it difficult to arrange home study visits. The wide geographical distribution of participants’ homes (30 – 140 miles from the research institute) restricted sample collection with a minimum of two hours required for sample collection and transportation to the laboratory.

We recruited controls from the same household as the patient to account for environmental confounders of microbiome analyses such as living conditions and diet^26, 27^. Previous studies have shown that inclusion of same household controls enables identification of ME/CFS disease-specific microbiome changes^28^. However, the requirement for a matched healthy household control impacted on patient recruitment. Amongst patients not meeting the study eligibility criteria, 44% were excluded due to being unable to identify a matched household control. Considering that household controls were typically carers and parents or spouses of the patient, it was not possible to match age and gender of patients and controls, both of which are confounding variables for immunological^29, 30^ and microbiome studies^31^. Other confounders potentially discordant amongst patients and controls were BMI, exercise and medications. Our finding of a sparse clustering between results from severe ME/CFS patients and matched household controls suggests that these and perhaps other confounding variables had an influence on immune response to the intestinal microbiome. Future studies should consider including and accounting for as many of these confounding variables in analysis as possible.

### Severe ME/CFS patients have serum IgG hyporeactivity to stool microbes

The hypothesis guiding our study was that ME/CFS patients have increased intestinal permeability due to an altered microbiome, which results in translocation of intestinal microbes into the circulation, triggering an immune response. Despite the low study sample size, we found evidence that severe ME/CFS patients in fact have lower serum IgG levels reactive to stool microbes than their matched household controls. This appears to be a property of the patients’ immune responses, rather than their microbiome which structurally and functionally closely resembled that of their matched controls. Our findings mirror the findings from pioneering studies investigating immune reactivity to faecal microbes in IBD patients which demonstrated that healthy controls have higher serum IgG reactive to heterologous stool microbes than to autologous stool microbes^15^. In our study heterologous reactivity was measured using samples from individuals living in the same household which are known to display greater similarity to patients’ microbiomes than individuals living in different households^32^. This should increase confidence in identifying microbes that distinguish the microbiome of patients from controls as being a property of the disease. Despite this advantage there were no differences in alpha diversity measures between severe ME/CFS patients and matched household controls at the genus or species level. Instead, our findings provide initial evidence for immune dysfunction in ME patients manifesting as a reduced capacity and reactivity of serum IgG to stool microbes irrespective of their source. Previous studies have described IgG immunodeficiencies in ME^33^ which could explain why we found reduced levels of serum IgG binding to stool microbes in severe ME/CFS patients. However, there was no evidence of IgG immunodeficiencies in our severe ME/CFS patient cohort. Instead, it may be attributable to IgG antibody repertoires and a less diverse repertoire of antibodies directed at intestinal microbes. The loss of antibody diversity occurs naturally during ageing and is a defining feature of immune senescence and declining immune function in later life^34^. The possibility of premature or accelerated immunosenescence should be a focus of future studies, particularly as it relates to effector immune cells whose functionality is compromised in ME patients, as previously described for NK cells^35^, and here for B cells. An additional possibility relates to immune hibernation in ME/CFS patients which produces a hypometabolic state^36^ that may limit lymphocyte responsiveness to foreign antigens and increased tolerance to bacterial endotoxins^37^.

### Limitations of the study in addressing the study hypothesis

The severe ME/CFS patients recruited to this pilot study had comorbid IBS, whereas household controls were free of any GI complaints. IBS comorbidity is a confounding variable in ME/CFS microbiome studies as defined microbiome profiles have been described that discriminate between patients with and without IBS comorbidity^9^. To establish whether the results found in this pilot study of severe ME patients are disease specific future studies should compare severe ME/CFS patients with and without comorbid IBS and compare severe ME/CFS patients with comorbid IBS to IBS patients.

We were unable to confirm the presence of increased intestinal permeability within the severe ME/CFS patient cohort. Based upon previous studies most ME patients (∼67%) will have increased intestinal permeability compared with healthy controls^13^. An intact intestinal barrier in the severe ME/CFS patients recruited to this study might explain why they did not have increased IgG reactivity to autologous stool bacteria compared with their matched healthy household controls. It has been shown that levels of IgG reactive to autologous stool bacteria are elevated in other diseases associated with increased intestinal permeability such as Crohn’s disease^16^. Future studies should seek to establish the integrity of the intestinal barrier in severe ME/CFS patients.

The IgG-Seq protocol used in this research identified serum IgG produced in response to bacteria and fungi. Whilst excluding reactivity to other constituents of the intestinal microbiome in the current study, the method can be adapted to detect IgG reactivity to viruses and archaea that can then be identified and isolated using FACS.

### Conclusion

Severe ME patients have historically been excluded from research studies. This pilot study demonstrates the feasibility and challenges associated with including this important and growing population of ME patients in research. In providing evidence of immune dysfunction in severe ME/CFS patients, expressed as hyporesponsive serum IgG responses to intestinal microbes, this study also provides the theoretical and methodological basis and rationale for undertaking more detailed immune function studies in larger cohorts of ME patients.

## Patients and Methods

### Participant recruitment

Ten volunteers were enrolled onto this study between 2018 and 2019: Five severe ME/CFS patients and, as controls, five healthy individuals that were the patients’ carers and living in the same household. Severe ME/CFS participants were recruited by the tertiary referral centres Epsom and St Helier University Hospitals (ESTH) CFS Service, Carshalton, UK, and the East Coast Community Healthcare Centre (ECCHC) ME/CFS service, Lowestoft, UK. All severe ME/CFS patients had a confirmed diagnosis of ME/CFS based on a hybrid of the NICE 2007 guidelines^38^ and the CDC-1994 criteria^39^ and defined as experiencing at least four of the following symptoms for a minimum of 4 months; cognitive difficulties, muscle pain, multi-joint pain, new headaches, recurrent sore throats, cervical/axillary lymphadenopathy, unrefreshing sleep and post exertional malaise. Severity was based on being unable to undertake activities associated with daily living, wheelchair dependency for mobility and being house- or bedbound and requiring aid for washing, using the toilet and eating. All patients were asked to complete the Chalder fatigue questionnaire, shortened medical outcomes study 36-item short form health survey (SF-36), hospital anxiety and depression scale (HADS), a self-efficacy questionnaire, visual analogue pain rating scale and the Epworth sleepiness scale. Matched household controls were defined as individuals who were living with or caring for the severe ME/CFS patient. Matched household controls were excluded if they had a long-term medical condition, in particular gastrointestinal conditions, autoimmune diseases, anxiety, or depression; or were receiving immunomodulatory drugs, statins, beta blockers or steroids. Participants consuming antibiotics or probiotic capsules within the 6 weeks prior to sample collection were excluded.

The study was performed in accordance with the Declaration of Helsinki and the International Conference on Harmonisation Good Clinical Practice (ICH GCP) Guideline, and in compliance with national law. The research was approved by the NHS Health Research Authority London Hampstead Research Ethics Committee (REC 17/LO/1102, IRAS ID 218545). This study was registered on the clinicaltrials.gov database (NCT03254823). All participants provided fully informed written consent. The collection, storage and use of human tissue samples was carried out within the terms of the Human Tissue Act 2004 (Human Tissue Authority).

### Sample collection and processing

Fresh stool and blood samples were collected from each participant within a 24 h window during a home visit. Twenty millilitres of blood were collected in serum separator tubes (BD Vacutainer). Serum was separated from whole blood following the manufacturers protocol and immediately stored at −80°C. Stool samples were collected immediately after defecation in a Fecotainer® (AT Medical BV) with an Oxoid™ AnaeroGen™ compact sachet (Thermo Scientific) to preserve anaerobic bacteria. The consistency and appearance of fresh stool samples was recorded using the Bristol stool form scale (BSFS) ^40^. Stool samples were stored at 4°C for <24 h. Following homogenisation aliquots of the bulk homogenate and of 40% stool microbial glycerol suspensions were stored at −80°C. Glycerol stocks were prepared by diluting stool samples (10% w/v) with PBS, collecting the supernatant following centrifugation at 300g for 5 min at 20°C and diluted 1:1 with 80% v/v glycerol.

### Stool water content

Non-diluted aliquots of stool were weighed before and after freeze drying using the ModulyoD freeze dryer (Thermo Electron Corp.) for 12 h. Water content was calculated using the following equation.

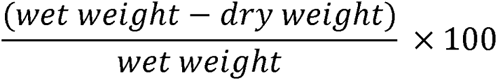

### Stool microbial load

Non-diluted stool aliquots were thawed on ice and diluted (1% w/v) with PBS solution containing 0.1% w/v BSA fraction V. Samples were homogenised using a Kimble™ Kontes™ Pellet Pestle™ Cordless motor and filtered through a 70 µm cell strainer. Filtered microbial suspensions were diluted 1:1600, 1:3200 and 1:6400; then 200µl of each dilution were incubated with 10µl of 1:100 SYBR™ Green I nucleic acid gel stain (Thermofischer Scientific) for 30 min. Microbial load was determined using the Guava® easyCyte™ 5HT equipped with a 488 nm laser. Prior to sample acquisition the instrument was cleaned following manufacturers’ instructions and calibrated using the Guava® easyCheck™ kit. Stool microbes (SYBR Green+ events) were enumerated using Guava® Suite Software v3.3.

### Stool IgA concentration

Non-diluted stool aliquots were thawed on ice, diluted (10% w/v) with 0.2M NaHCO_3_, pH 9.4 and homogenised using a Kimble™ Kontes™ Pellet Pestle™ Cordless motor. Samples were centrifuged twice at 16,000g for 5 min at 20°C and after both rounds of centrifugation the pellet and supernatant were separated. The supernatants were pooled and used to analyse free secretory IgA (sIgA) by ELISA. The pellet was washed and resuspended to the original volume and used to analyse microbe associated/bound sIgA by ELISA. All reactants except for the blocking solution and wash buffers were added in volumes of 100 µl/well. All washes were repeated three times with PBS and 0.05% Tween™ 20 unless otherwise stated. Nunc MaxiSorp™ flat-bottom plates (Thermo Fisher Scientific) were coated for 16h at 4°C with 10-fold serial dilutions (1 to 1:1000000) of samples and with 2-fold serial dilutions (250-3.9 ng/ml) of a human colostrum IgA standard (Sigma), diluted with 0.2M NaHCO_3_, pH 9.4. Samples and standards were plated in duplicate. Plates were washed and then blocked for 3h with 300µl/well of PBS, 0.05% Tween™ 20, 2% BSA fraction V, 1% normal mouse serum (Thermo Fisher Scientific). Plates were washed and then incubated with 1000 ng/ml biotin conjugated mouse anti-human IgA1/IgA2 monoclonal antibody (Pharmingen) diluted in PBS, 0.05% Tween™ 20, 2% BSA fraction V, 1% normal mouse serum. After 1h of incubation at 20°C plates were washed and incubated with 1:80000 HRP-Conjugated Streptavidin (Thermo Fisher Scientific) diluted in PBS, 0.05% Tween™ 20, 2% BSA fraction V, 1% normal mouse serum for 30 min at 20°C. Plates were washed six times and incubated with TMB high sensitivity substrate solution (BioLegend® UK Ltd) for 5 min. The addition of 2N H_2_SO_4_ stopped the reaction and absorbances were read at 450 nm. IgA sample values were determined by reference to standard curves.

### Serum IgG quantification

Total serum IgG was measured using the commercial Invitrogen ELISA kit (Thermo Fisher Scientific). Samples were measured in duplicate.

### Serum IgG levels to autologous and heterologous stool microbes

Glycerol stocks of stool microbes were thawed and washed three times with PBS at 8000g for 5 min at 20°C. The pellet was resuspended in 1ml PBS. Serial dilutions (2-fold) of stool microbes in PBS were plated in duplicate in a 96-well flat bottom Corning™ Costar™ 9018 plate. Absorbances were read at 570 nm and stool microbes were resuspended to an optical density of 0.05 with 0.1M NaHCO_3_, pH 9.4.

Serum IgG reactivity to autologous (‘self’) or heterologous (‘non-self’, matched patient or control) stool microbes was measured by ELISA. All washes were repeated three times with 200 µl/well PBS and 0.1% Tween™ 20 unless otherwise stated. Nunc MaxiSorp™ flat-bottom plates (Thermo Fisher Scientific) were coated for 16h at 4°C with 100 µl/well of stool microbes. Plates were washed and then blocked for 3h at room temperature under agitation with PBS, 2% BSA fraction V, 1% normal goat serum (Sigma-Aldrich). Plates were incubated for 1 h at 20°C with 50 µl/well of complement inactivated serum samples 2-fold serially diluted (1:80-1:320) in PBS, 2% BSA fraction V, 1% normal goat serum. Plates were washed and incubated for 1h at room temperature under agitation with 100 µl/well of 1:500 goat anti-human IgG H&L (HRP) (Abcam,) diluted in PBS, 2% BSA fraction V, 1% normal goat serum. Plates were washed six times and incubated with TMB high sensitivity substrate solution (BioLegend® UK Ltd) for 5 min at 20°C. The addition of 0.16M H_2_SO_4_ stopped the reaction and absorbances were read at 450 nm. Results were normalised by subtracting serum only absorbance readings from sample readings.

### Microbial flow cytometry

Systemic IgG and faecal IgA binding to stool microbes was assessed by microbial flow cytometry. All buffers were filter sterilised using a 0.22-µm filter before use. Aliquots of non-diluted stool samples were processed and the microbial concentration in each stool sample measured as described earlier. Microbes were resuspended to 2 x 10^6^ cells/ml in PBS with 0.1% BSA. 500µl of complement inactivated serum was diluted 1:100 in PBS with 0.1% BSA and incubated for 30 min at 20°C with 200 µl of 1 x 10^6^ cells/ml of stool microbes. For secretory IgA measurements stool microbes were not incubated with serum. Samples were washed in PBS with 0.1% BSA (5 min, 1.2 x 10^4^ rpm) and microbes were resuspended to 1 x 10^6^ cells/ml and incubated with 1:1000 SYBR™ Green I nucleic acid gel stain and 1:100 secondary conjugated antibodies (anti-human IgA-APC (Miltenyi Biotec) and anti-human IgG-APC/Cy7 (BioLegend) or their respective isotype controls. Samples were fixed with 0.75% v/v paraformaldehyde. Acquisition of cellular events was performed using the BD LSRFortessa™ (BD Biosciences) and analysed using FlowJo™ software. Frequencies of antibody bound stool microbes were expressed as percentages. The percentage of Ig-bound microbes was normalised by subtracting frequency of Ig-bound microbes measured using isotype controls.

### Metagenomic shotgun sequencing analysis of total gut microbiota and IgG-coated microbes (IgG-Seq)

#### Microbial cell sorting

Non-diluted stool samples were processed and stained for IgG flow cytometry analysis as described previously. Isotype controls were not used in microbial cell sorting. Microbes were diluted to 1 x 10^7^ cells/ml and cell sorting achieved using the Sony SH800S cell sorter equipped with four lasers: 488 nm, 405nm, 638nm and 651nm. Prior to sample acquisition the instrument was cleaned and calibrated according to the manufacturer’s instructions. Microbes (1 x 10^6^) were collected from the following three fractions: 1) ‘all’ (SYBR^+^ microbes), 2) ‘IgG positive’ (SYBR^+^IgG^+^), 3) ‘IgG negative’ (SYBR^+^IgG^-^). Fractions were centrifuged and immediately stored at −20°C as dry pellets.

#### DNA extraction, processing and sequencing

DNA was extracted using the Gram-positive bacteria genomic DNA purification protocol from the GeneJET DNA genomic DNA purification kit (Thermoscientific) with the following modifications: 1) 0.52 kU/ml achromopeptidase was added to the lysis buffer and the incubation time increased to 60 min; 2) incubation with lysis solution and proteinase K was increased to 50 min. DNA was precipitated using 0.7X solid phase reversible immobilisation bead clean-up with KAPA pure beads (Roche). Whole genome amplification was performed using the REPLI-g advanced DNA single cell kit (QIAGEN). The quality and quantity of amplified genomic DNA were determined using the Agilent Tapestation 4200 and the Quanti-iT™ dsDNA high sensitivity assay kit (Thermo Fisher Scientific). A ready-to-load pooled sequencing library was prepared by the in-house QIB sequencing service using the Illumina DNA prep kit (Illumina, 20018704) and the KAP2G Robust PCR kit (Sigma), followed by sequencing using 2 x 150 bp paired-end chemistry (PE150) on the Illumina NovaSeq 6000 platform (Novogene Ltd., UK).

Paired-end sequencing reads were provided as FASTQ format. All raw sequencing reads were pre-processed using tools retrieved from the BioConda repository^41^. SeqFu (v1.8.5) ^42^ was used to assess the quality of raw sequencing reads and those bases below Phred quality score of 15 were removed using Fastp (v0.20.0) ^43^. Human genomic DNA identified by Kraken2^44^ against the Genome Reference Consortium Human Build 37 (GRCh37/hg19) database were removed. Taxonomic assignment of remaining sequencing reads was done using Kraken2^44^ against the ‘PlusPF’ database containing archaea, bacteria, viral, plasma, human1, UniVec_Core, Protozoa and Fungi https://benlangmead.github.io/aws-indexes/k2. Abundance of reads at the species level was estimated using Bracken^45^. Taxonomic read counts were converted to relative abundances by total sum scaling to 1.

#### Relative and quantitative microbiome profiling

The ‘all’ fraction from microbial cell sorting was used for relative microbiome profiling (RMP) and quantitative microbiome profiling (QMP). The cut off threshold of 1 x 10^-6^ was applied to relative abundances and a pseudocount of 1 x 10^-7^ was added. The microbial load of each species was calculated on these modified relative abundances using the equation below.

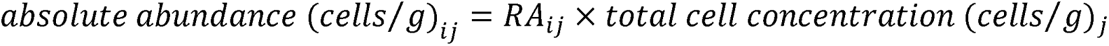

#### Analysing IgG binding of taxa

RMP data was used for IgG binding analysis with a cut off threshold of 1 x 10^-5^. In addition, for each participant if a species was not detected in the ‘all’ fraction above the threshold then this species was subsequently removed from respective ‘IgG positive’ and ‘IgG negative’ fractions. The ‘IgAScores’ (v.0.1.2) ^25^ R package was used to calculate IgG probability ratios using the ‘igascores’ function with method set to ‘probratio’, pseudocount ‘c’ set to 1 x 10^-5^ and ‘scaleratio’ set to TRUE.

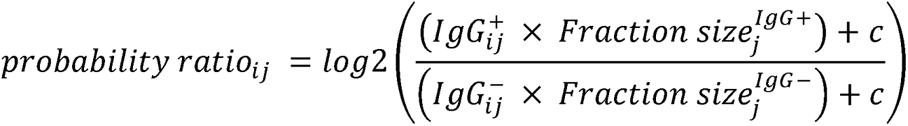

#### Alpha diversity

The diversity function from the ‘vegan’ R package (v.2.5-7) ^46^ was used to calculate Shannon indices and inverse Simpson indices. The rarefy function from ‘vegan’ (v.2.5-7) ^46^ was used to rarefy reads to the lowest sequencing depth. Observed species’ richness was the number of species remaining following rarefaction.

#### Beta diversity

The vegdist function from ‘vegan’ (v.2.5-7) ^46^ was used to calculate Bray-Curtis indices on relative abundances. The metMDS function from ‘vegan’ (v.2.5-7) ^46^ was used to performed non-metric multidimensional scaling (NDMS) on Bray-Curtis indices.

#### Functional analysis

Gene families were identified using the HMP Unified Metabolic Analysis Network 3.0 (HUMAnN 3.0) package from the bioBakery suite^47, 48^. In all participants each gene family was filtered by the following criteria: if a gene family was not present in the ‘all’ fraction the reads per kilobase (RPK) were zeroed in the ‘IgG positive’ and ‘IgG negative’ fractions of that participant. The humann_renorm_table utility script from HUMAnN 3.0 was used to convert gene families from RPK to relative abundances. Community level classifications of gene families were used in downstream analysis. A cut off threshold of 1 x 10^-6^ was applied to all samples.

To analyse gene families in the microbiome of severe ME/CFS patients compared with household controls the ‘all’ fraction was used. Gene families that were below the threshold in seven or more samples were discarded from downstream analysis. The clr function from the ‘compositions’ R package (v 2.0-4) ^49^ was used to CLR transform the relative abundance of gene families. The pca function from the ‘mixOmics’ R package (v 6.18.1) ^50^ was used to perform principal component analysis (PCA) on gene families. Severe ME/CFS patients and their matched household controls were not treated as paired samples in this analysis.

To analyse gene families of IgG reactive microbes IgG probability ratios were calculated using the relative abundance of gene families in the ‘IgG positive’ fraction and the ‘IgG negative’ fraction as described earlier, with a pseudocount set to 6 x ^-^10. Gene families that did not have IgG probability ratios in all samples were discarded from downstream analysis. PCA was then performed as described previously.

### Statistical analysis

Statistical analyses and graphical representations were performed in R using the following packages: ‘ggplot2’ (v3.4.0) ^51^, ‘reshape2’ (v1.4.4) ^52^, ‘data.table’ (v1.14) ^53^, ‘dplyr’ (v1.1.0) ^54^, ‘ggpubr’ (V0.5.0) ^55^. Graphs were also made using GraphPad Prism 5.04. Prior to analysis data were log-transformed if there was evidence for non-normality. Pairwise comparisons between severe ME/CFS patients and their matched household controls were done using a two-tailed paired *t*-test. Correlations were assessed with Pearson (*r*) correlation test.

### Common data elements for ME research

This study used the National Institute of Neurological Disorders and Stroke Common Data Elements guidelines’ for reporting microbiome/microorganisms biomarkers in ME/CFS research (http://www.commondataelements.ninds.nih.gov/)^56^.

## Data Availability

The raw WMS data are available at the European Nucleotide Archive under study accession PRJEB61661.

http://www.ebi.ac.uk/ena/broswer/view/PRJEB61661

## Acknowledgements

We acknowledge East Coast Community Healthcare ME/CFS service and the CFS clinic at Epsom and St Helier University Hospitals NHS Trust for their help recruiting patients, the QIB Human Research Governance Committee (HRGC) for reviewing the HRA application and all the research participants who generously shared their time. KAS was supported by a PhD studentship jointly funded by Invest in ME Research (UK charity number 1153730) and the University of East Anglia. The authors gratefully acknowledge the support of the Biotechnology and Biological Sciences Research Council (BBSRC); The authors gratefully acknowledge the support of the Biotechnology and Biological Sciences Research Council (BBSRC); this research was supported by the BBSRC Institute Strategic Programme Grant Gut Microbes and Health BB/R012490/1 and its constituent projects BBS/E/F/000PR10353 and BBS/E/F/000PR10355 (SRC). MD and GS were funded by the BBSRC Core Capability Grant BB/CCG1860/1.

## Data availability statement

The raw WMS data are available at the European Nucleotide Archive under study accession PRJEB61661. (http://www.ebi.ac.uk/ena/broswer/view/PRJEB61661).

---

**SUPPLEMENTARY TABLE 1.** The size of IgG positive and IgG negative fractions collected during IgG-Seq. This data was used to calculation of IgG probability ratio scores.

| Pair | Participant | Fraction sizes (%) |  |
| --- | --- | --- | --- |
|  |  | IgG+ | IgG- |
| 1 | Patient | 21.1 | 79.5 |
|  | Control | 18.0 | 79.8 |
| 2 | Patient | 35.1 | 58.1 |
|  | Control | 48.1 | 49.7 |
| 3 | Patient | 17.9 | 65.5 |
|  | Control | 46.3 | 44.9 |
| 4 | Patient | 65.0 | 29.0 |
|  | Control | 33.7 | 63.1 |
| 5 | Patient | 24.2 | 75.3 |
|  | Control | 62.7 | 31.1 |

SUPPLEMENTARY FIGURE 1. Study participant recruitment pathway. Severe ME patients were recruited from the CFS clinic at Epsom and St Helier University Hospitals (ESTH), Carshalton, UK and the ME/CFS service at East Coast Community Healthcare Centre, Lowestoft, UK.

SUPPLEMENTARY FIGURE 2. Stool microbial load. Flow cytometric analysis of SYBR Green+ microbial load in stool samples of severe ME/CFS patients (n=5) and matched household controls (n=5).

SUPPLEMENTARY FIGURE 3. Stool microbiome profiling of severe ME/CFS patients (n=5) and matched household controls (n=5). (A) Relative microbiome profiling (RMP) at the genus-level. (B) quantitative microbiome profiling (QMP, cells per gram of faeces) at the genus-level. (C) RMP at the species-level. (D) QMP at the species-level.

SUPPLEMENTARY FIGURE 4. Pairwise alpha- and beta-diversity comparisons of the stool microbiomes of severe ME/CFS patients (n=5) and matched household controls (n=5). Analyses were performed at the species-level on shotgun metagenomics data from SYBR+ stool microbes. (A) Alpha diversity measures of Shannon index, inverse Simpson index and richness based on reads rarefied to the lowest sequencing depth. P values were measured using two-tailed paired t-tests. (B) Beta diversity of relative microbiome profiling (RMP) and quantitative microbiome profiling (QMP, cells per gram faeces). Beta-diversity was calculated using Bray-Curtis dissimilarity, presented on a non-metric multi-dimensional scaling (NMDS) plot.

SUPPLEMENTARY FIGURE 5. Functional composition of stool metagenomes from severe ME/CFS patients (circles) (n=5) and matched household controls (squares) (n=5). Principal component analysis (PCA) of the relative abundances of gene families. Pair numbers are depicted on the graph but were not used in analysis.

SUPPLEMENTARY FIGURE 6. Functional profiling of stool microbes reactive with serum IgG. Principal component analysis (PCA) of IgG probability ratios of gene families from the stool microbiome in severe ME/CFS patients (circles) (n=5) and matched household controls (squares) (n=5). Pair numbers are depicted on the graphs but were not used in analysis.

