## Supplementary figures and images for "Investigating antibody reactivity to the intestinal microbiome in severe myalgic encephalomyelitis/chronic fatigue syndrome (ME/CFS)"

### Fig. S1

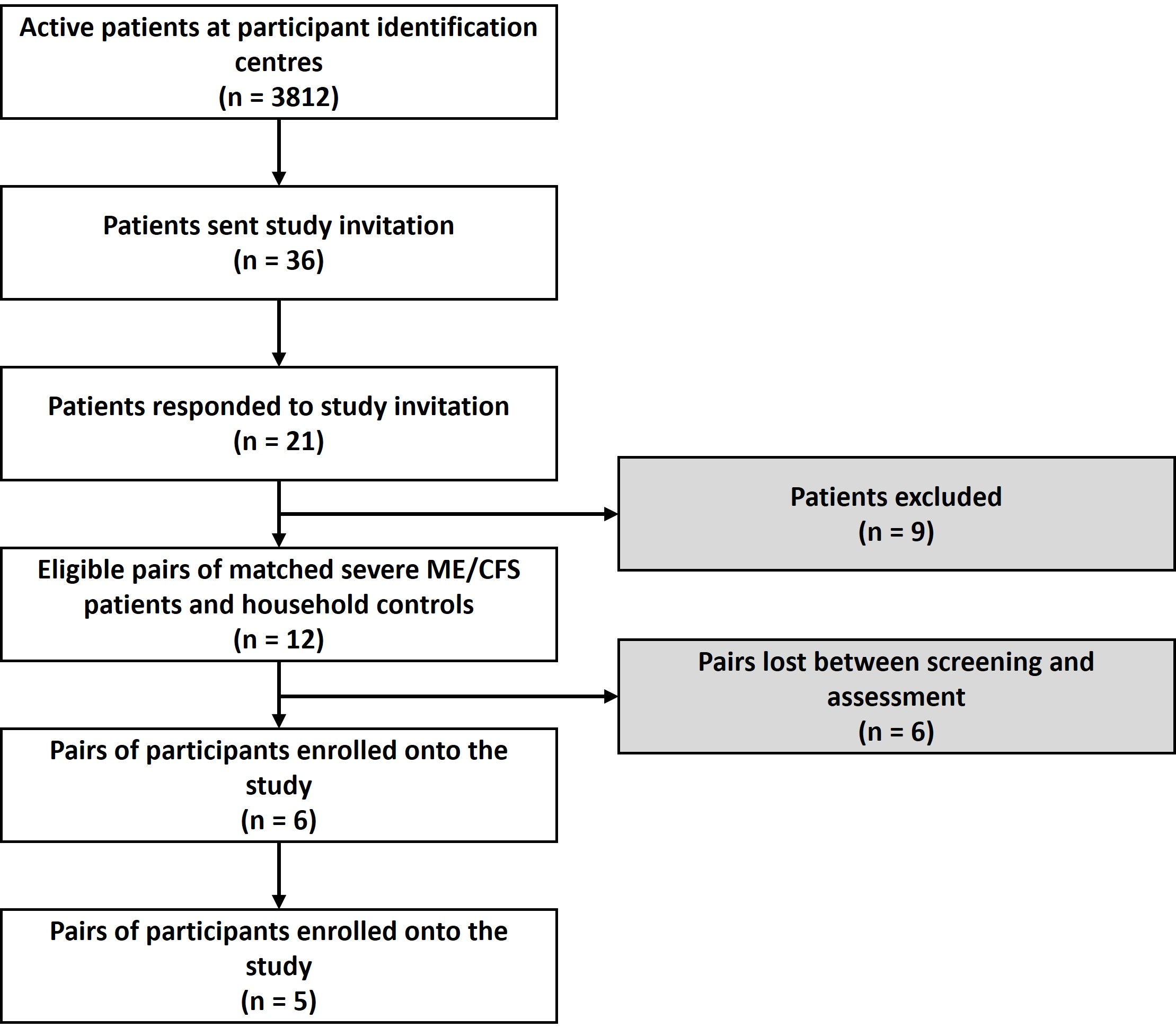

### Fig. S2

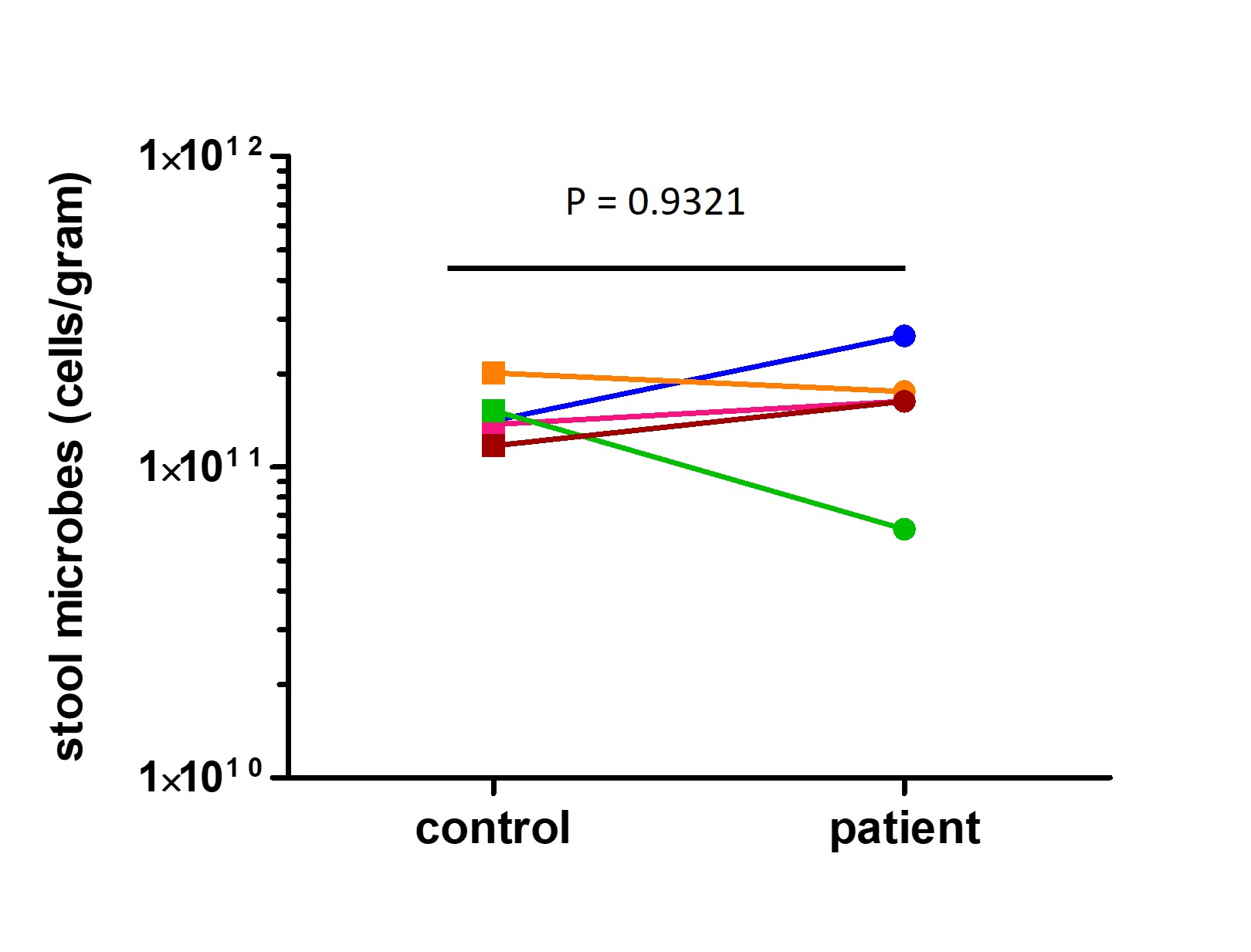

### Fig. S3

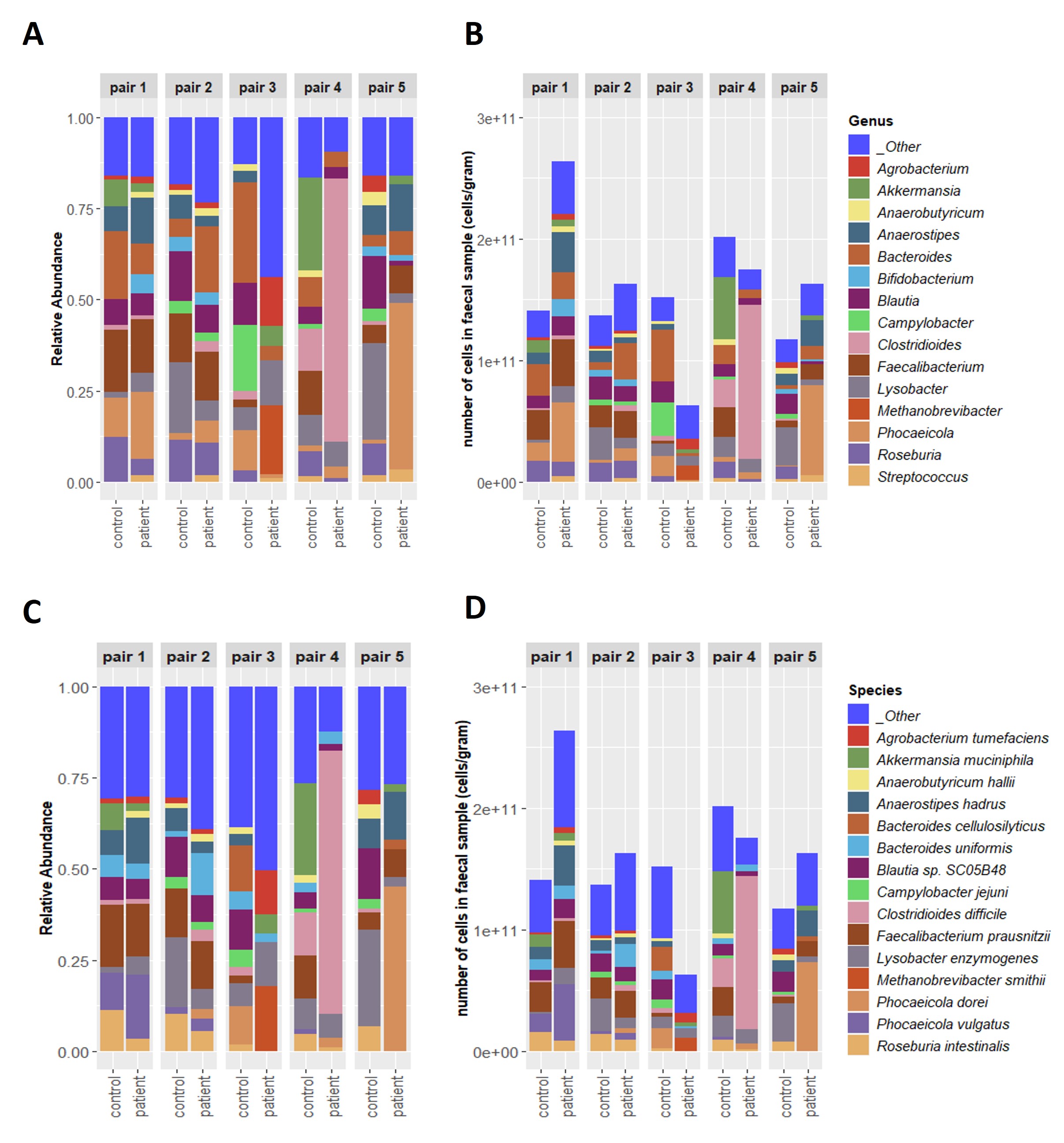

### Fig. S4

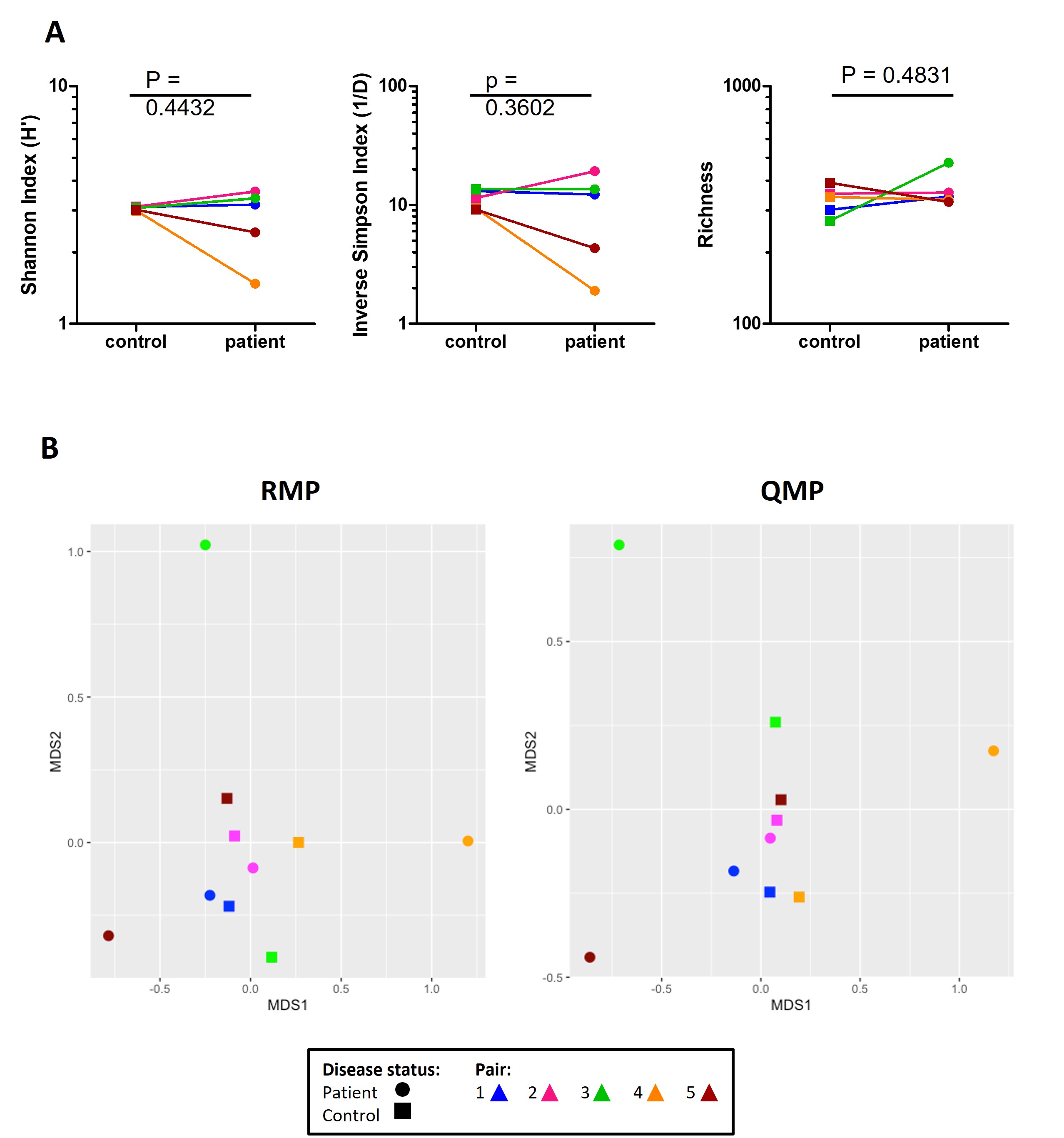

### Fig. S5

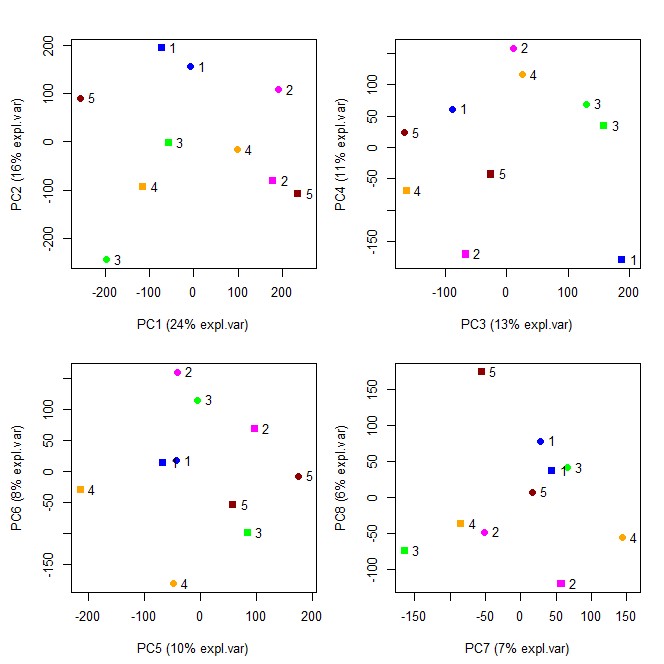

### Fig. S6

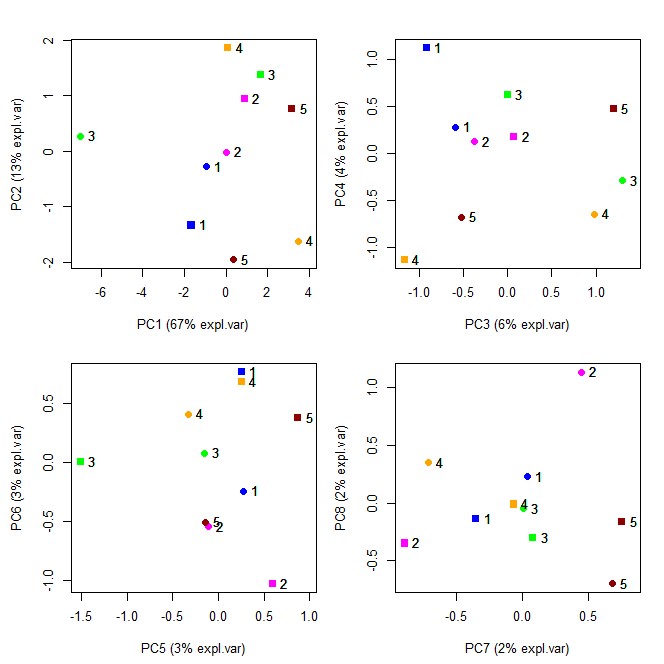
